# Evaluation of the Anticipated Burden of COVID-19 on Hospital-Based Healthcare Services Across the United States

**DOI:** 10.1101/2020.04.01.20050492

**Authors:** Rohan Khera, Snigdha Jain, Zhenqiu Lin, Joseph S. Ross, Harlan M Krumholz

## Abstract

**Background:** Coronavirus disease-19 (COVID-19) is a global pandemic, with the potential to infect nearly 60% of the population. The anticipated spread of the virus requires an urgent appraisal of the capacity of US healthcare services and the identification of states most vulnerable to exceeding their capacity

**Methods:** In the American Hospital Association survey for 2018, a database of US community hospitals, we identified total inpatient beds, adult intensive care unit (ICU) beds, and airborne isolation rooms across all hospitals in each state of continental US. The burden of COVID-19 hospitalizations was estimated based on a median hospitalization duration of 12 days and was evaluated for a 30-day reporting period.

**Results:** At 5155 US community hospitals across 48 states in the contiguous US and Washington DC, there were a total of 788,032 inpatient beds, 68,280 adult ICU beds, and 44,222 isolation rooms. The median daily bed occupancy was 62.8% (IQR 58.1%, 66.6%) across states. Nationally, for every 10,000 individuals, there are 24.2 inpatient beds, 2.8 adult ICU beds, and 1.4 isolation beds. There is a 3-fold variation in the number of inpatient beds available across the US, ranging from 16.4 per 10,000 in Oregon to 47 per 10,000 in South Dakota. There was also a similar 3-fold variation in available or non-occupied beds, ranging from 4.7 per 10,000 in Connecticut through 18.3 per 10,000 in North Dakota. The availability of ICU beds is low nationally, ranging from 1.4 per 10,000 in Nevada to 4.7 per 10000 in Washington DC.

Hospitalizations for COVID-19 in a median 0.2% (IQR 0.2 %, 0.3%) of state population, or 1.4% of state’s older adults (1.0%, 1.9%) will require all non-occupied beds. Further, a median 0.6% (0.5%, 0.8%) of state population, or 3.9% (3.1%, 4.6%) of older individuals would require 100% of inpatient beds.

**Conclusion:** The COVID-19 pandemic is likely to overwhelm the limited number of inpatient and ICU beds for the US population. Hospitals in half of US states would exceed capacity if less than 0.2% of the state population requires hospitalization in any given month.

## Background

Coronavirus disease-19 (COVID-19) is a global pandemic, with the potential to infect nearly 60% of the population.^1^ The disease is associated with high morbidity and mortality and current estimates are that nearly 20% of patients require hospitalization, 5% critical care.^2,3^ Over the past 2 months, multiple countries have witnessed their healthcare systems being overwhelmed by patients infected by COVID-19. The first case was diagnosed in the United States on January 21, 2020, but it has spread rapidly across the US.^4,5^ The anticipated spread of the virus requires an urgent appraisal of the capacity of US healthcare services and the identification of states most vulnerable to exceeding their capacity.

## Methods

We evaluated state-level variation in bed capacity and ICU bed availability, and the state population requiring hospitalization that would saturate healthcare services. In the American Hospital Association survey for 2018, a database of US community hospitals, we identified total inpatient beds, adult intensive care unit (ICU) beds, and airborne isolation rooms across all hospitals in each state of continental US. The bed occupancy rate was defined as the daily average patient-days per hospital bed. We normalized our estimates against 2018 US Census estimates. The burden of COVID-19 hospitalizations was estimated based on a median hospitalization duration of 12 days,^3^ and was evaluated for a 30-day reporting period. All statistics are descriptive and were calculated using Stata 16.

## Results

In 2018, there were 5155 US community hospitals across 48 states in contiguous US and Washington DC, with a total of 788,032 inpatient beds, 68,280 adult ICU beds, and 44,222 isolation rooms. The median daily bed occupancy was 62.8% (IQR 58.1%, 66.6%) across states. Nationally, for every 10,000 individuals, there are 24.2 inpatient beds, 2.8 adult ICU beds, and 1.4 isolation beds.

There is a 3-fold variation in the number of inpatient beds available across the US, ranging from 16.4 per 10,000 in Oregon to 47 per 10,000 in South Dakota (**Figure 1**). There was also a similar 3-fold variation in available or non-occupied beds, ranging from 4.7 per 10,000 in Connecticut through 18.3 per 10,000 in North Dakota. The availability of ICU beds is low nationally, ranging from 1.4 per 10,000 in Nevada to 4.7 per 10000 in Washington DC.

**Figure 1:**
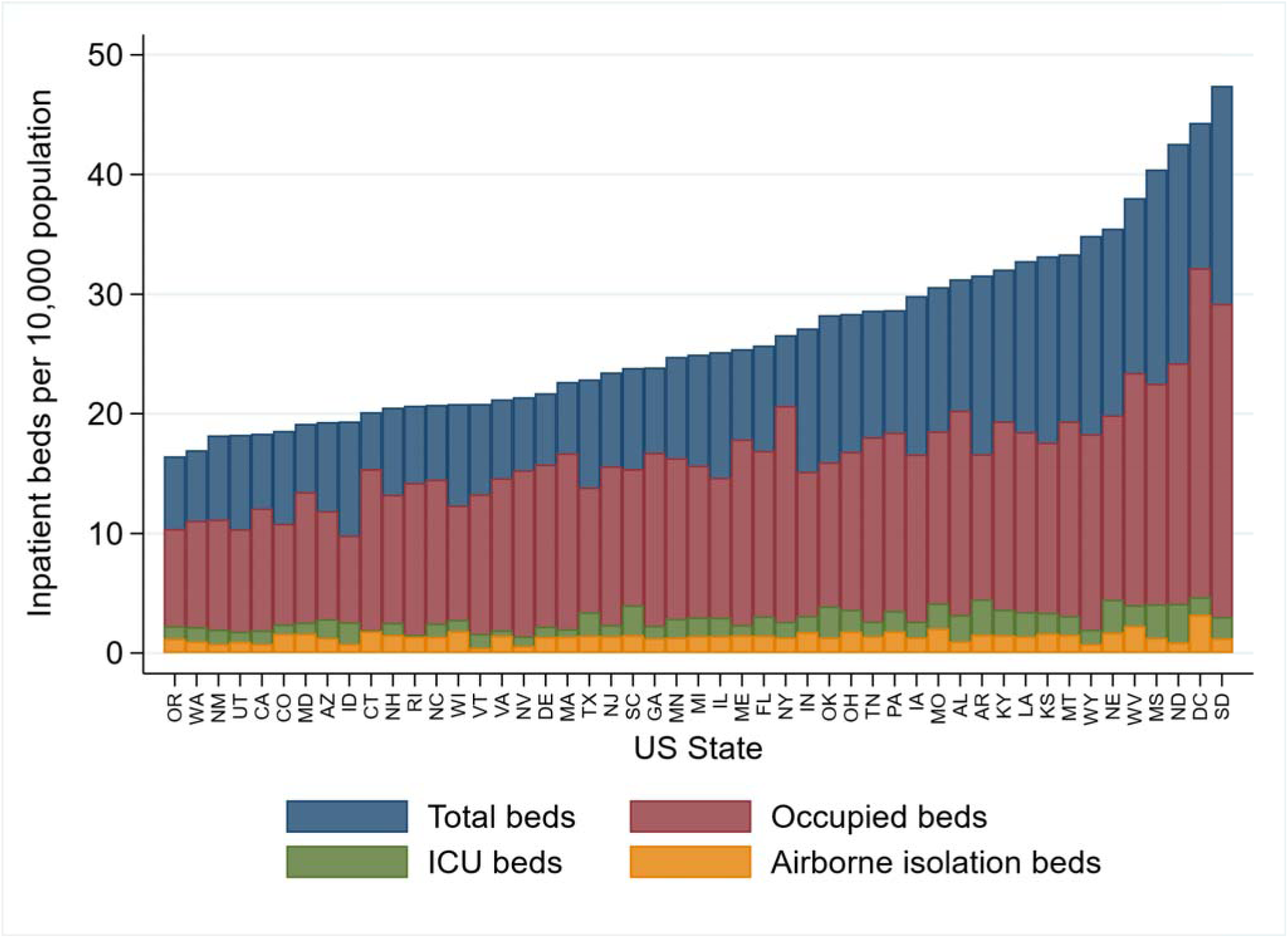
Inpatient bed capacity across US states. Number of inpatient beds per 10,000 population in 2018, overall (blue), occupied (based on state-wise occupancy rates), intensive care unit (ICU, green) and airborne isolation (yellow).

Hospitalizations for COVID-19 in a median 0.2% (IQR 0.2 %, 0.3%) of state population (Figure 2), or 1.4% of state’s older adults (1.0%, 1.9%) will require all non- occupied beds. Further, a median 0.6% (0.5%, 0.8%) of state population, or 3.9% (3.1%, 4.6%) of older individuals would require 100% of inpatient beds. States of Vermont, Wyoming, Delaware, and Rhode Island would exceed their capacity with 2000 hospitalizations in a month, while Texas, California and Florida can each accommodate nearly 50 thousand patients in their available capacity.

**Figure 2:**
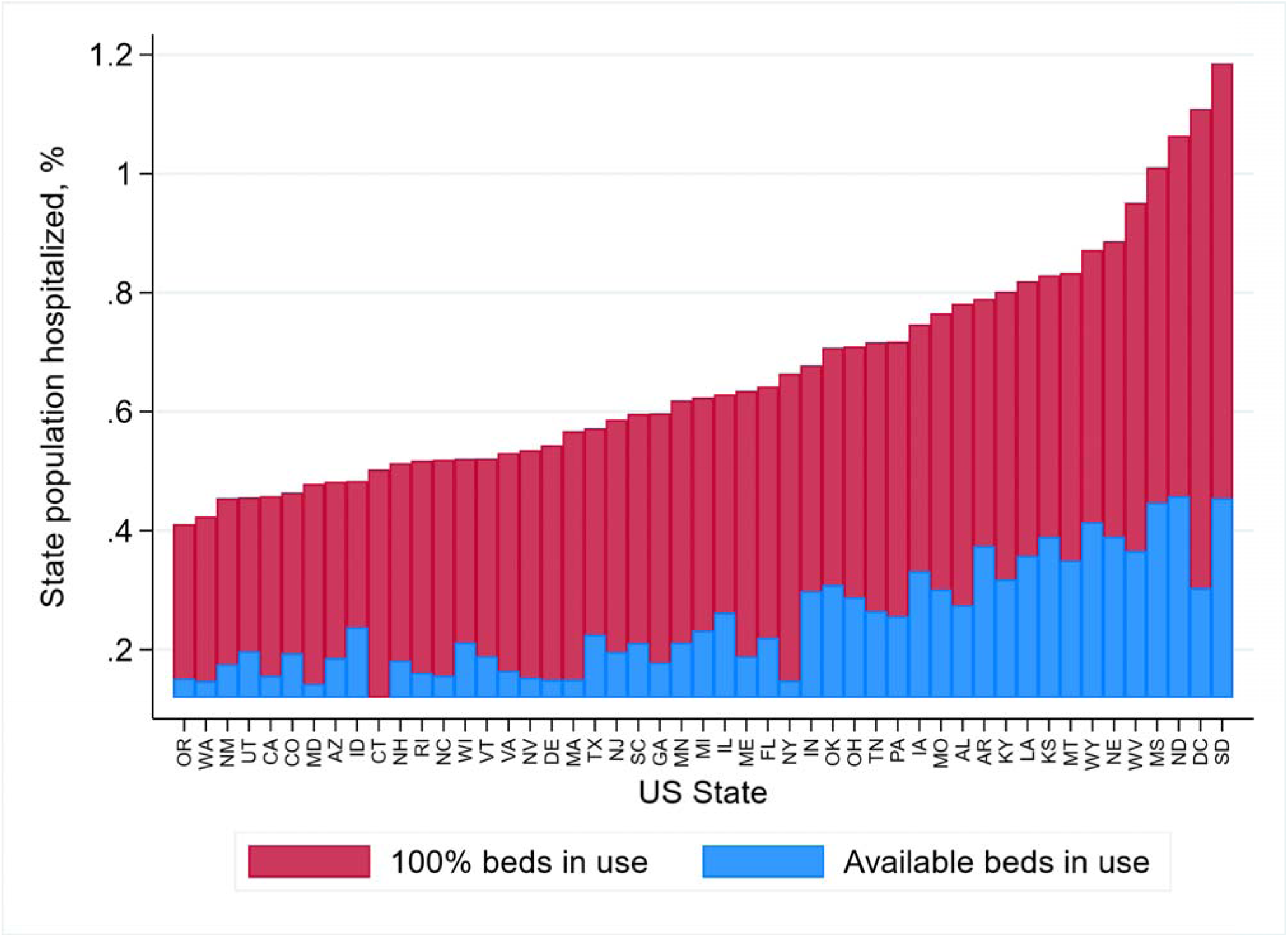
State monthly hospitalization rates and hospital capacity. The percentage of a state’s population that can be hospitalized with COVID-19 over a 30-day period before all inpatient beds based on 2018 hospital utilization rates (all available beds in blue) and if 100% of a state’s inpatient beds (in red) are used.

## Discussion

The COVID-19 pandemic is likely to overwhelm the limited number of inpatient and ICU beds for the US population. Hospitals in half of US states would exceed capacity if less than 0.2% of state population requires hospitalization in any given month. These are equivalent to rates observed in Wuhan, China, 1% of the population was affected, 1 in 5 of whom required hospitalization.^2^

The study has certain limitations. We assume that all beds can be used to care for patients infected by COVID-19, however, expected concurrent needs for other disease conditions and long-term healthcare needs among those with a chronic course would suggest that our estimates are conservative.

There is an urgent need to deploy epidemiological strategies to contain the spread of COVID-19,^6^ with a focus on states with limited healthcare capacity. Concurrently, we may need to expand capacity quickly, possibly through suspension of elective hospitalizations.

## Data Availability

Data in the report are proprietary of the American Hospital Association.

